# The Role of Inflammation in Depression: A Scoping Review Protocol in Mechanisms, Evidence, and Therapeutic Potential

**DOI:** 10.1101/2025.01.05.25320002

**Authors:** Yan Bo

## Abstract

**Introduction:** Depression is a leading global cause of disability and premature mortality, yet a substantial proportion of patients do not respond adequately to standard treatments. An increasing body of evidence suggests that inflammatory processes contribute to the onset, persistence, and recurrence of depression; however, this evidence remains fragmented across basic, translational, and clinical research domains.

**Methods and analysis:** This scoping review aims to systematically map the existing evidence on the role of inflammation in depression and to characterise how inflammatory markers and pathways have been investigated in relation to prevention and treatment strategies. The review will be conducted in accordance with established methodological guidance for scoping reviews and reported following the Preferred Reporting Items for Systematic Reviews and Meta-Analyses extension for Scoping Reviews (PRISMA-ScR). Multiple bibliographic databases, trial registries, grey literature sources, and preprint servers will be searched without language restrictions. Two reviewers will independently screen records, select eligible studies, and chart data using a piloted data extraction form. Findings will be synthesised using descriptive statistics and narrative methods to summarise study characteristics, inflammatory mechanisms, biomarkers, interventions, and clinical outcomes. An integrative framework will be developed to link inflammatory mechanisms with therapeutic and preventive approaches.

**Ethics and dissemination:** This review will use only publicly available data, and no new data will be collected; therefore, formal ethical approval is not required. The findings will be disseminated through peer-reviewed publication and other academic dissemination channels to support knowledge translation in depression research.

**Registration:** This scoping review protocol is registered with the Open Science Framework (DOI: https://doi.org/10.17605/OSF.IO/SE49K).

**Strengths and limitations of this study:** *Strengths:*

▪ This scoping review provides a comprehensive synthesis of mechanistic evidence examining the role of inflammation in depression, integrating findings across clinical, preclinical, and translational research.
▪ The inclusion of grey literature and preprints enhances the breadth of evidence captured and reduces the risk of publication bias.
▪ Transparency and reproducibility are strengthened through prospective protocol registration, adherence to established scoping review guidelines, and peer review of the research process.

*Limitations:*

▪ As a scoping review, this study does not undertake formal critical appraisal of individual studies or quantitative synthesis of effect sizes, which limits causal inference.
▪ The heterogeneity of study designs, populations, and inflammatory measures may constrain direct comparison across studies.
▪ Reliance on published and publicly available sources may result in incomplete capture of unpublished or ongoing research.

## 1 Introduction

Depression represents a substantial global disease burden [1]. According to the World Health Organization (WHO), more than 700,000 individuals with depression die by suicide each year, and as of May 2023 an estimated 280 million people worldwide were affected by the disorder [2]. Despite extensive reporting of preventive strategies and therapeutic interventions, no country has succeeded in eliminating the disease burden associated with depression. Affected individuals experience marked reductions in personal productivity, alongside impairments in quality of life and social functioning [3,4]. At present, advocacy for the implementation and dissemination of evidence-based treatment strategies remains limited [5]. From a biological perspective, this may partly reflect the absence of sufficiently convincing mechanistic explanations underlying the effectiveness of these interventions [6]. From a sociological perspective, inadequate public awareness and promotion of available treatments may further contribute to this gap [7].

Existing evidence broadly categorises interventions for depression into two principal approaches: pharmacological and non-pharmacological. However, no single intervention has been shown to be unequivocally superior in terms of overall effectiveness [7,8]. Pharmacological treatments are commonly associated with limited durability of therapeutic benefit, substantial financial costs, and a considerable burden of adverse effects [9]. By contrast, non-pharmacological interventions often involve prolonged treatment courses, high demands on human resources, and suboptimal levels of acceptability among target populations [10,11]. It has been reported with a high degree of confidence that, on average, the equivalent human resources of eight trained professionals are required to successfully prevent one case of depression [7]. Nevertheless, preventive intervention strategies have been shown to reduce the incidence of major depressive disorder by approximately 25–50% [12].

These findings suggest that prevention-oriented approaches may play a pivotal role in future efforts to reduce the population-level burden of depression. In this context, a global transnational project on depression prevention, initiated in Utrecht, the Netherlands, in September 2011, was associated not only with improvements in mental health outcomes but also with reductions in the incidence and mortality of cardiovascular disease [7,13].

An increasing body of evidence has documented an association between inflammation and depression in recent years [14–16], stimulating interest in the development of inflammation-targeted treatment strategies for depressive disorders [16,17]. This emerging literature provides a biological rationale for certain non-pharmacological interventions, particularly physiotherapy-based approaches, and offers a novel perspective on evidence-based treatment paradigms for depression.

The present hypothesis seeks to synthesise existing evidence into a coherent theoretical framework for the inflammatory hypothesis of depression, grounded in established anti-inflammatory biological mechanisms [18,19]. Such a synthesis may help to address current gaps in the dissemination and promotion of evidence-based treatment strategies. This hypothesis is aligned with prevailing clinical guidelines and with the core objective of scoping reviews, namely the systematic mapping, synthesis, and dissemination of existing evidence [20,21]. Accordingly, this study aims to comprehensively synthesise the evidence on the role of inflammation in depression, following established methodological guidelines for scoping reviews.

This scoping review aims to systematically integrate and organise the existing evidence underpinning the inflammatory hypothesis of depression.

## 2 Methods and analysis

### 2.1 Design

This study is designed as a scoping review and will be reported in accordance with the Preferred Reporting Items for Systematic Reviews and Meta-Analyses extension for Scoping Reviews (PRISMA-ScR) [22]. The review protocol and any subsequent updates are registered and publicly available on the Open Science Framework (OSF) [23].

The review will follow the established methodological framework for scoping reviews, originally proposed by Arksey and O’Malley [20] and subsequently refined by Levac et al. [21] and the Joanna Briggs Institute [22]. Scoping reviews are a form of evidence synthesis that systematically map existing literature and summarise the current state of research within a given field [24]. In contrast to systematic reviews, scoping reviews do not aim to generate pooled effect estimates, which makes them particularly suitable for examining broad, heterogeneous, or conceptually complex research questions [25–28].

The scoping review process comprises six key stages, which are described in detail below: (1) identifying the research question; (2) identifying relevant studies; (3) study selection; (4) data charting; (5) collating, summarising, and reporting the results; and (6) consultation and dissemination.

### 2.2 Patient and public involvement

As this study is a retrospective evidence synthesis based exclusively on previously published literature, all data analysed were collected during the original studies. Consequently, patients and members of the public were not directly involved in the design of the study protocol or in the data collection processes of this review. This complies with the data security and legal requirements as set out by Yifei Chen when establishing the research plan[29].

### 2.3 Eligibility criteria

As summarised in **Table 1**, the eligibility criteria were formulated using the Population, Intervention, Comparator, and Outcomes (PICO) framework [30]. **Table 1** presents the detailed inclusion and exclusion criteria applied in this review.

**Table 1.**
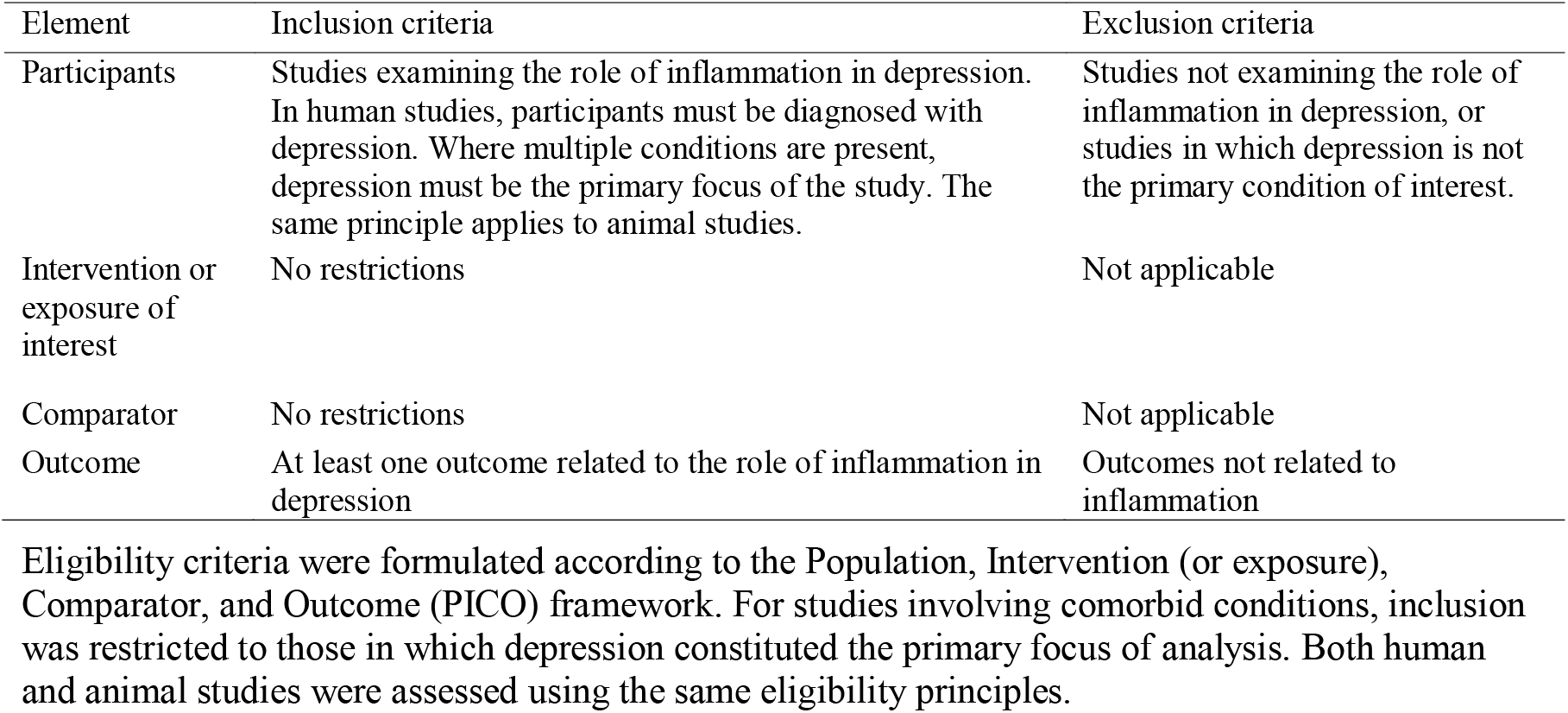
Eligibility criteria based on the PICO framework.

| Element | Inclusion criteria | Exclusion criteria |
| --- | --- | --- |
| Participants | Studies examining the role of inflammation in depression. In human studies, participants must be diagnosed with depression. Where multiple conditions are present, depression must be the primary focus of the study. The same principle applies to animal studies. | Studies not examining the role of inflammation in depression, or studies in which depression is not the primary condition of interest. |
| Intervention or exposure of interest | No restrictions | Not applicable |
| Comparator | No restrictions | Not applicable |
| Outcome | At least one outcome related to the role of inflammation in depression | Outcomes not related to inflammation |

Eligibility criteria were formulated according to the Population, Intervention (or exposure), Comparator, and Outcome (PICO) framework. For studies involving comorbid conditions, inclusion was restricted to those in which depression constituted the primary focus of analysis. Both human and animal studies were assessed using the same eligibility principles.

In this scoping review, two broad categories of literature will be included. First, primary studies, comprising clinical trials, cohort studies, case–control studies, and other original research designs, which provide primary data on the association between inflammation and depression. Second, evidence synthesis studies, including systematic reviews and scoping reviews, which facilitate the identification of research trends, knowledge gaps, and conceptual developments within the field, thereby offering a broader contextual perspective for evidence synthesis.

During data extraction and synthesis, primary studies and evidence synthesis studies will be analysed and reported separately. Primary studies will be used to characterise study-specific designs, populations, and findings, whereas evidence synthesis studies will be examined to appraise the consistency and divergence of existing evidence and to inform directions for future research.

### 2.4 Step 1: Identifying the research question

The research questions for this scoping review were developed by a multidisciplinary team with expertise in evidence-based medicine, emergency medicine, psychiatry, public health, and basic biomedical research. Consistent with established methodological guidance for scoping reviews, the overarching objective was to identify and structure a coherent body of evidence within a large and heterogeneous literature base that addresses a population-level health problem of substantial public relevance [20,21].

The primary research question guiding this scoping review was: **What role does inflammation play in depression?**

For the purposes of this review, depression was defined in accordance with the diagnostic criteria outlined in the Diagnostic and Statistical Manual of Mental Disorders, Fifth Edition (DSM-5) [31]. The concept of inflammation was operationally defined based on Medical Subject Headings (MeSH) terminology in PubMed as a pathological process involving tissue injury and immune-mediated cellular and molecular responses, encompassing both acute and chronic inflammatory states.

As emphasised by Levac et al., the formulation of research questions in scoping reviews should be closely aligned with the intended purpose of the review and the anticipated outcomes of evidence synthesis [21]. Accordingly, the objective of the present scoping review is to provide a comprehensive overview of the existing evidence on the role of inflammation in depression. The resulting synthesis is intended to inform and refine a conceptual framework for understanding the inflammatory hypothesis of depression.

To further delineate the scope of the review, a set of secondary research questions was developed to guide study identification, data extraction, and synthesis:

1. Which inflammatory biomarkers or indicators are associated with the development of depression?
2. How do varying levels or patterns of inflammation differentially influence depressive symptoms or phenotypes?
3. What biological pathways have been proposed to link inflammation with the onset or progression of depression?
4. What roles do specific inflammation-related cytokines play in depressive disorders?
5. What clinical characteristics are observed in patients with comorbid inflammatory conditions and depression?
6. How is the effectiveness of anti-inflammatory interventions evaluated in the context of depression?
7. How do lifestyle and environmental factors modify the relationship between inflammation and depression?
8. How does the association between inflammation and depression vary across different age groups?
9. Are there sex-specific differences in the relationship between inflammation and depression?
10. What evidence exists regarding the effectiveness of psychological interventions for inflammation-associated depression?

### 2.5 Step 2: Identifying relevant studies

A comprehensive yet pragmatic search strategy will be implemented to ensure the systematic identification of all relevant literature. Electronic searches will be conducted across multiple databases, with strategies tailored to the specific indexing structure of each database. The search process will be designed and executed by researchers with expertise in evidence-based medicine, using structured database-specific frameworks to maximise sensitivity and coverage.

The core search strategy will be developed in PubMed using Medical Subject Headings (MeSH) and corresponding entry terms, and subsequently adapted for use in other databases. The full PubMed search strategy is provided in the **Supplementary Materials**. No restrictions will be applied with respect to publication date or language in order to capture the broadest possible range of potentially relevant studies. Studies lacking sufficient detail to address the objectives of this scoping review will be excluded during the screening process.

Consistent with methodological guidance for scoping reviews, the search strategy will be iterative, with refinements made as necessary to improve the identification of relevant evidence. All modifications to the search strategy will be documented and reported through updates to the review protocol. In addition, backward and forward citation tracking will be undertaken to identify further eligible studies, following approaches commonly employed in evidence-based medicine research [32–35].

References cited in relevant systematic reviews, including topic-specific meta-analyses and systematic evaluations, will also be examined to minimise the risk of missing pertinent literature.

To ensure timely access to emerging evidence, preprint servers will be searched as an extension of the primary strategy. This approach aims to capture recent research outputs that may not yet be indexed in traditional databases. Preprint sources will include arXiv, bioRxiv, F1000, Figshare, PeerJ Preprints, Zenodo, Oxford University ePrints, Nature Precedings, and medRxiv.

Grey literature will be identified using a structured approach informed by the grey literature search checklist and related guidance published by the Canadian Agency for Drugs and Technologies in Health (CADTH) [36,37]. The grey literature search strategy will be developed through a stepwise exploratory process led by researchers trained in evidence-based medicine.

### 2.6 Step 3: Study selection

Arksey and O’Malley recommend the explicit specification of inclusion and exclusion criteria when conducting a scoping review [20]. Building on this approach, Levac et al. further emphasise the importance of ongoing team discussions to refine eligibility criteria throughout the study selection process [21]. In accordance with this guidance, initial eligibility criteria were developed a priori, as described below. These criteria were subsequently refined through iterative team meetings, with any substantive modifications documented through updates to the review protocol.

The eligibility framework was structured hierarchically, progressing from the population or phenomenon of interest, to study design, and finally to study content. Specifically, studies were first assessed for relevance to the research focus; only those meeting this initial criterion were then evaluated with respect to study type and substantive content.

#### 2.6.1 Identification and removal of duplicate records

Following retrieval of records from all databases, all references will be imported into the Zotero reference management software. Duplicate records will be identified and removed using Zotero’s built-in de-duplication (merge) function. This automated process will be used to ensure consistency and efficiency in duplicate removal, without the need for manual screening at this stage.

#### 2.6.2 Title and abstract screening

As this review includes both primary studies and evidence synthesis studies, a structured, multi-stage screening process will be employed. The screening procedure will comprise four sequential stages: (1) title and abstract screening, (2) full-text retrieval, (3) full-text eligibility assessment, and (4) final report selection.

Prior to formal screening, two independent reviewers will pilot the screening process by jointly assessing a random sample of 50 records. Screening will proceed once an inter-reviewer agreement of at least 80% is achieved, after which dual independent screening will be conducted for all remaining records.

During title and abstract screening, studies will be required to have depression as the primary focus, without restriction to specific depressive subtypes, such as treatment-resistant depression, bipolar disorder, or major depressive disorder. Eligible records must examine at least one aspect of the association between inflammation and depression. For evidence synthesis studies, including systematic reviews, the article must explicitly address a conceptual or theoretical perspective on the relationship between inflammation and depression.

At this stage, records will be categorised as either primary research or evidence synthesis studies. This classification will be used to inform subsequent stages of screening and data extraction..

#### 2.6.3 Full-text screening and eligibility assessment

##### (1) Full-text screening

During the full-text screening stage, a secondary eligibility assessment will be conducted to confirm alignment with the core focus of the review. Consistent with methodological guidance for evidence synthesis, priority will be given to information presented in sections with greater methodological weight, including the Methods and Results, followed by the Introduction, Discussion, and Conclusions, with the Abstract assigned the lowest priority [38]. Where inconsistencies arise across sections within a single report, eligibility decisions will be based on the section with the highest priority, in order to minimise misclassification due to superficial or ambiguous descriptions.

Studies will be required to explicitly state that the research involved human participants, or to clearly define an objective examining the relationship between inflammation and depression. The definition of depression will be standardised according to the diagnostic criteria outlined in the Diagnostic and Statistical Manual of Mental Disorders, Fifth Edition (DSM-5) [31], with eligibility restricted to studies in which depression is the primary condition under investigation. This approach is consistent with established practices in prior systematic reviews employing comparable selection strategies[39,40].

Studies focusing on broader clinical entities in which depression is not the principal condition of interest will be excluded. Examples of excluded study contexts include:

a. studies primarily examining medical conditions (e.g. heart failure) with depression reported only as a secondary outcome;
b. studies addressing mixed psychiatric phenotypes, such as anxiety–depression comorbidity, without a distinct analytical focus on depression.

##### (2) Type of study

Methodological guidance in evidence-based research has highlighted that the exclusive inclusion of randomised controlled trials may enhance the internal validity of systematic reviews [41]. However, scoping reviews are designed to accommodate a broader range of study designs in order to comprehensively map existing evidence across a field.

Accordingly, this scoping review will include original empirical studies employing both observational and experimental designs. Eligible observational studies will include case reports, cohort studies, case–control studies, and cross-sectional studies, while experimental studies will include randomised controlled trials [42] (**Table 1**). All included primary studies must present a clearly described Methods section detailing the study design and analytical approach.

Evidence synthesis studies, such as systematic reviews and scoping reviews, will not be included as sources of primary data but will be used to contextualise findings and inform interpretation in the Discussion section.

##### (3) Outcomes of interest and data prioritisation

As shown in **Table 1**, direct outcome definitions related to inflammation were not uniformly reported across the included literature. To address this heterogeneity, study eligibility at the outcome level was guided by the ten secondary research questions defined a priori. To be included, each study was required to address at least one of these secondary questions. This structured approach to outcome identification is conceptually aligned with the analytical strategies used in prior systematic reviews addressing complex and multifaceted research topics [39].

During full-text assessment, each eligible article was examined systematically against the secondary research questions, with particular attention to conceptual clarity, operational definitions, and methodological relevance. This process facilitated consistent identification of outcomes related to the role of inflammation in depression across diverse study designs.

##### (4) Study screening and reviewer agreement

Title/abstract screening and full-text screening were conducted independently by two reviewers working in parallel. Discrepancies were resolved through discussion, and when consensus could not be reached, a third reviewer was consulted to make a final determination.

Prior to formal screening, a pilot assessment was undertaken to evaluate inter-reviewer agreement. Screening commenced once a satisfactory level of agreement was achieved (Cohen’s κ ≥ 0.60), consistent with commonly accepted methodological thresholds for evidence synthesis. Conference abstracts, correspondence, and publications unrelated to the research topic were excluded. For studies published in languages unfamiliar to the reviewers, preliminary assessment was supported using automated translation tools.

##### (5) Full-text assessment and data prioritisation

Following retrieval of full-text articles, reviewers applied differential screening priorities according to study type.

For original human studies, priority was given to outcomes including validated depression symptom scales (e.g. SDS, HAMD, MADRS), inflammatory biomarker concentrations, depression-associated neurotransmitters (e.g. serotonin, norepinephrine, γ-aminobutyric acid), and indicators of neuroimmune activity (e.g. CD4□ and CD8□ T-cell subsets).

For basic or preclinical studies involving animal models, cellular systems, or molecular analyses, screening focused on depression-like behavioural phenotypes and neuropathological correlates relevant to depressive disorders.

For evidence synthesis studies, assessment emphasised conceptual integration, identification of knowledge gaps, and the contribution of novel interpretative frameworks. To fall within scope, such studies were required to address both inflammation and depression explicitly.

References from eligible review articles were further examined using backward citation tracking to identify potentially relevant primary studies.

##### (6) Data management, transparency, and documentation

All records were managed using Zotero reference management software, with duplicate records removed using automated de-duplication procedures. Decisions made during full-text screening, including reasons for exclusion, were documented to ensure transparency and reproducibility.

Extracted data and methodological updates will be deposited on the Open Science Framework (OSF) [23]. The review process adheres to principles of transparency and replicability, enabling independent verification and secondary analyses where appropriate.

### 2.7 Step 4: Charting the Data

To facilitate systematic data extraction, the review team collaboratively developed a standardised data charting form (**Table 2**). This form was designed to ensure consistent and comprehensive capture of relevant information across studies and was informed by established reporting guidance, including the Guidelines for Reporting on Patient and Public Involvement (GRIPP2) checklist [43]. Reviewers were trained to ensure a shared understanding of each data item prior to formal extraction.

**Table 2.**
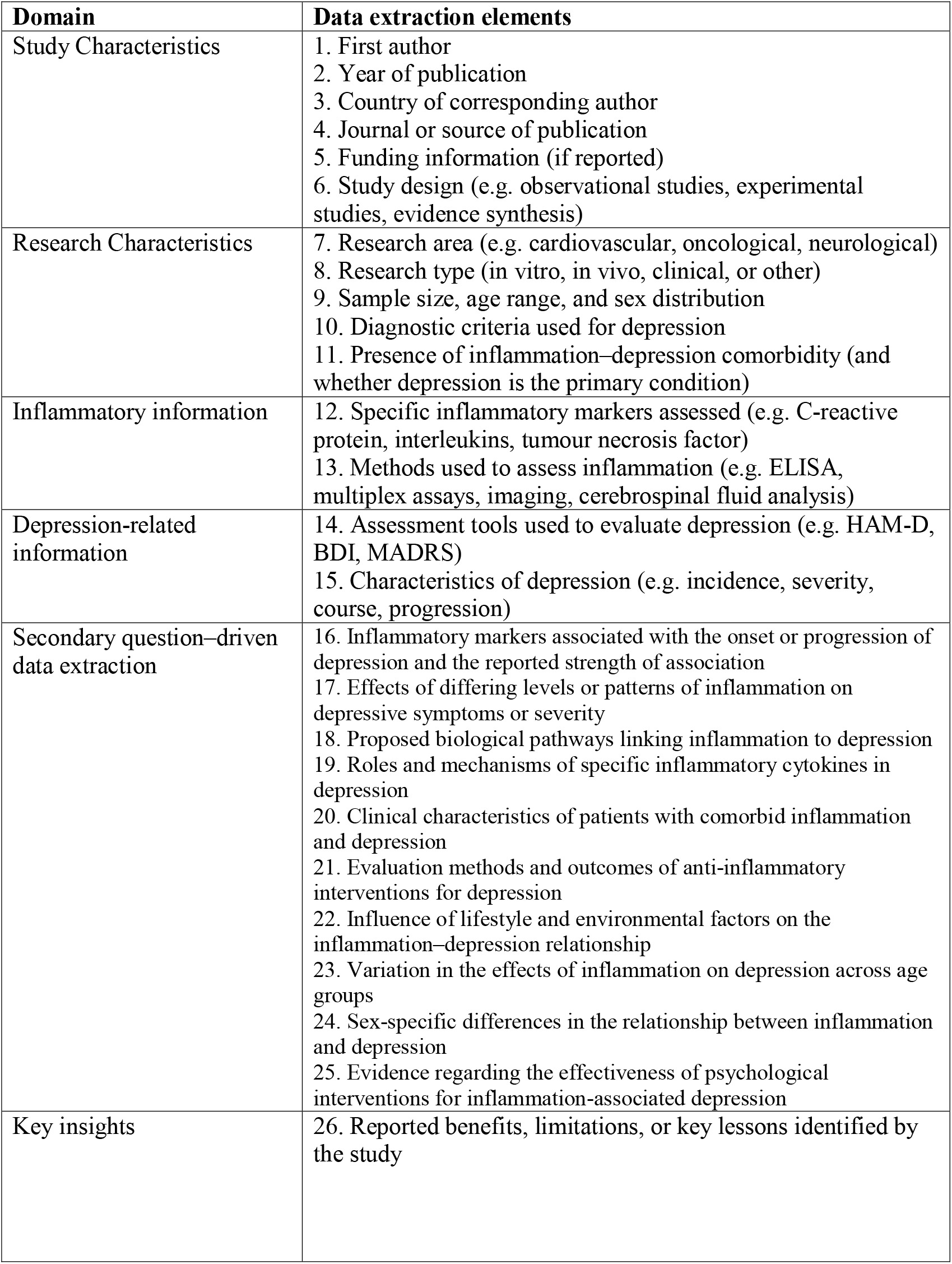
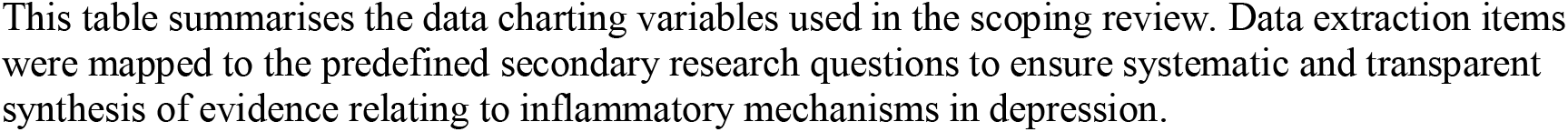
Data charting items and variables extracted in the scoping review.

| **Domain** | **Data extraction elements** |
| --- | --- |
| Study Characteristics | 1. First author<br>2. Year of publication<br>3. Country of corresponding author<br>4. Journal or source of publication<br>5. Funding information (if reported)<br>6. Study design (e.g. observational studies, experimental studies, evidence synthesis) |
| Research Characteristics | 7. Research area (e.g. cardiovascular, oncological, neurological)<br>8. Research type (in vitro, in vivo, clinical, or other)<br>9. Sample size, age range, and sex distribution<br>10. Diagnostic criteria used for depression<br>11. Presence of inflammation–depression comorbidity (and whether depression is the primary condition) |
| Inflammatory information | 12. Specific inflammatory markers assessed (e.g. C-reactive protein, interleukins, tumour necrosis factor)<br>13. Methods used to assess inflammation (e.g. ELISA, multiplex assays, imaging, cerebrospinal fluid analysis) |
| Depression-related information | 14. Assessment tools used to evaluate depression (e.g. HAM-D, BDI, MADRS)<br>15. Characteristics of depression (e.g. incidence, severity, course, progression) |
| Secondary question–driven data extraction | 16. Inflammatory markers associated with the onset or progression of depression and the reported strength of association<br>17. Effects of differing levels or patterns of inflammation on depressive symptoms or severity<br>18. Proposed biological pathways linking inflammation to depression<br>19. Roles and mechanisms of specific inflammatory cytokines in depression<br>20. Clinical characteristics of patients with comorbid inflammation and depression<br>21. Evaluation methods and outcomes of anti-inflammatory interventions for depression<br>22. Influence of lifestyle and environmental factors on the inflammation–depression relationship<br>23. Variation in the effects of inflammation on depression across age groups<br>24. Sex-specific differences in the relationship between inflammation and depression<br>25. Evidence regarding the effectiveness of psychological interventions for inflammation-associated depression |
| Key insights | 26. Reported benefits, limitations, or key lessons identified by the study |

This table summarises the data charting variables used in the scoping review. Data extraction items were mapped to the predefined secondary research questions to ensure systematic and transparent synthesis of evidence relating to inflammatory mechanisms in depression.

In accordance with methodological recommendations for scoping reviews, the data charting form will be iteratively refined as reviewers become increasingly familiar with the included literature [21]. A pilot data extraction exercise will be conducted on a subset of approximately ten studies, and inter-reviewer consistency will be assessed using Cohen’s kappa coefficient, as recommended by the Joanna Briggs Institute [22]. Any modifications to the data charting form arising from this process will be documented.

Where essential information is missing or unclear, attempts will be made to contact the corresponding authors for clarification.

#### 2.7.1 Data domains and outcomes of interest

Data extraction will focus on characterising the relationship between inflammation and depressive symptoms, with particular attention to disease course and treatment response. The following outcome domains will be extracted where reported:

1. Clinical rating scales: Validated depression severity measures, including the Hamilton Depression Rating Scale (HAM-D) and the Beck Depression Inventory (BDI), will be extracted to quantify depressive symptom burden.
2. Neuroimaging intermediate phenotypes: Neuroimaging data, including functional magnetic resonance imaging (fMRI) and structural magnetic resonance imaging (sMRI), will be extracted where available to examine associations between inflammation and alterations in brain structure or function relevant to depression.
3. Inflammatory markers: Peripheral inflammatory biomarkers commonly reported in depression research will be extracted, including C-reactive protein (CRP), interleukin-6 (IL-6), and tumour necrosis factor-α (TNF-α), as well as other cytokines where relevant.
4. Quality of life outcomes: Measures of health-related quality of life, such as the Short Form-36 (SF-36) or the World Health Organization Quality of Life (WHOQOL) instruments, will be extracted to capture functional outcomes beyond core depressive symptoms.

#### 2.7.3 Peripheral and central inflammatory markers

For peripheral inflammation, data will be extracted on major inflammatory markers, including CRP, IL-6, TNF-α, and other cytokines, together with the biological specimens analysed and measurement methods reported (e.g. enzyme-linked immunosorbent assay [ELISA], bead-based multiplex assays).

For central nervous system inflammation, cerebrospinal fluid (CSF) biomarkers, such as IL-6, TNF-α, and interleukin-1β (IL-1β), will be extracted where available, along with the corresponding analytical techniques (e.g. ELISA or flow cytometry).

During data charting, peripheral and central inflammatory markers will be clearly distinguished, and details of assay methods will be recorded to facilitate comparability and interpretability across studies.

### 2.8 Step 5: Collating, summarizing and reporting the results

The results of this scoping review will be collated and synthesised to provide an integrated overview of the existing evidence on the role of inflammation in depression. Quantitative data will be summarised using descriptive statistics to characterise study features and research trends, with findings presented in tabular and graphical formats to facilitate intuitive interpretation of the current evidence landscape.

Qualitative data extracted from textual sources will be analysed using an inductive thematic analysis approach [44,45]. This process will be used to synthesise definitions, identify similarities and differences across studies, and highlight knowledge gaps related to inflammatory mechanisms in depression. It is particularly important to emphasise here that in the process of integrating textual evidence, we may utilise certain special algorithmic components, such as the Newton downhill optimiser. This specific algorithmic component currently holds some application value in cancer-related fields[46].

Textual data will be independently coded by two reviewers. An initial coding framework will be developed to organise and describe key concepts emerging from the data. Discrepancies in coding will be resolved through discussion, and the coding framework will be iteratively refined as necessary.

Codes exhibiting conceptual similarity will be grouped into descriptive themes to capture recurring patterns across the evidence base. These themes will be tabulated to allow comparison within and between studies, with rows representing individual studies and columns representing thematic domains [44,45]. Where possible, descriptive summaries will be used to illustrate convergence and divergence in findings.

The identified themes will subsequently be reviewed by the full research team to support the development of higher-order analytical themes. These analytical themes will be synthesised into a narrative summary explaining how inflammatory processes have been conceptualised and investigated in relation to depression.

Finally, drawing on the integrated quantitative and qualitative findings, a conceptual framework will be developed to summarise the proposed mechanisms linking inflammation and depression. This framework will be refined iteratively as themes are reviewed and consolidated, with the aim of informing future research directions and the development of inflammation-informed approaches to depression research.

### 2.9 Dissemination of the scoping review findings

The findings of this scoping review will be disseminated to ensure broad accessibility and effective knowledge translation. Primary dissemination will occur through peer-reviewed academic publication, allowing the results to contribute to the existing scholarly literature on inflammation and depression.

In addition, the review findings will be communicated through publicly accessible academic and educational formats, including invited lectures and online presentations, to enhance understanding among clinical, academic, and general audiences. To facilitate accessibility for non-specialist readers, simplified visual materials, such as brief animated summaries, will also be developed and shared via internet-based platforms and social media.

The dissemination of knowledge is intended to be achieved through the utilisation of snowballing statistical principles [47]. These dissemination activities are intended solely to communicate the results of the scoping review and do not constitute additional data collection or influence the evidence synthesis process.

## Data Availability

All data produced in the present work are contained in the manuscript.

https://osf.io/2phwv/

## 3 Project timeline

The anticipated timeline for the scoping review is outlined below.

Research protocol publication: from January 2025 to January 2026

Literature search: from 1 January 2026 to 8 January 2026

Literature screening: from 8 January 2026 to 8 April 2026

Data extraction: from 8 April 2026 to 8 August 2026

Data synthesis and analysis: from 8 August 2026 to 8 October 2026

Manuscript drafting and revision: from 8 October 2026 to 8 January 2027

## 4 Ethics

This scoping review did not require additional ethical approval, as it was based exclusively on data obtained from publicly available databases and grey literature, all of which had received ethical approval in their original studies where applicable.

The findings of this review will be disseminated through peer-reviewed publication in the public domain. In accordance with open-access principles, the results will be made available under a Creative Commons licence to facilitate access by individuals with depression, clinicians, and researchers.

## Declaration

### AI disclosure

The authors declare that no Gen AI was used in the creation of this manuscript.

### Author contributions

(CRediT): The author approved the final version of the manuscript and agrees to be accountable for all aspects of the work.YB: Conceptualization; Investigation; Data curation; Formal analysis; Resources; Visualization; Writing – original draft; Writing – review & editing; Supervision; Project administration.

## Acknowledgements

The study has been registered on the Open Science Framework (OSF) platform at the following address: osf.io/2phwv. Registration DOI: https://doi.org/10.17605/OSF.IO/SE49K

## Acronyms

PRESS: peer review of electronic search strategies
PRISMA: preferred reporting64 items for systematic reviews and meta-analyses
PRISMA-ScR: preferred reporting65 items for systematic reviews and meta-analyses extension for scoping reviews
OSF: open science framework
CADTH: Canadian Agency for Drugs and Technologies in Health
PICO: participants, intervention,comparator and outcomes.

## References

1. Global burden of 369 diseases and injuries in 204 countries and territories, 1990-2019: a systematic analysis for the Global Burden of Disease Study 2019. Lancet. 2020;396:1204–22. doi: 10.1016/S0140-6736(20)30925-9

2. Depressive disorder (depression). World Health Organization. 2023. https://www.who.int/news-room/fact-sheets/detail/depression (accessed 3 January 2025)

3. Saxena S, Funk M, Chisholm D. World Health Assembly adopts Comprehensive Mental Health Action Plan 2013-2020. Lancet. 2013;381:1970–1. doi: 10.1016/S0140-6736(13)61139-3

4. Brundtland GH. From the World Health Organization. Mental health: new understanding, new hope. JAMA. 2001;286:2391. doi: 10.1001/jama.286.19.2391

5. Thornicroft G, Alem A, Antunes Dos Santos R, et al. WPA guidance on steps, obstacles and mistakes to avoid in the implementation of community mental health care. World Psychiatry. 2010;9:67–77. doi: 10.1002/j.2051-5545.2010.tb00276.x

6. Insel TR. The NIMH Research Domain Criteria (RDoC) Project: precision medicine for psychiatry. Am J Psychiatry. 2014;171:395–7. doi: 10.1176/appi.ajp.2014.14020138

7. Cuijpers P, Beekman ATF, Reynolds CF 3rd. Preventing depression: a global priority. JAMA. 2012;307:1033–4. doi: 10.1001/jama.2012.271

8. Cipriani A, Furukawa TA, Salanti G, et al. Comparative efficacy and acceptability of 21 antidepressant drugs for the acute treatment of adults with major depressive disorder: a systematic review and network meta-analysis. Lancet. 2018;391:1357–66. doi: 10.1016/S0140-6736(17)32802-7

9. Gartlehner G, Hansen RA, Morgan LC, et al. Comparative benefits and harms of second-generation antidepressants for treating major depressive disorder: an updated meta-analysis. Ann Intern Med. 2011;155:772–85. doi: 10.7326/0003-4819-155-11-201112060-00009

10. Cuijpers P, Karyotaki E, Pot AM, et al. Managing depression in older age: psychological interventions. Maturitas. 2014;79:160–9. doi: 10.1016/j.maturitas.2014.05.027

11. Cuijpers P, Donker T, Weissman MM, et al. Interpersonal Psychotherapy for Mental Health Problems: A Comprehensive Meta-Analysis. Am J Psychiatry. 2016;173:680–7. doi: 10.1176/appi.ajp.2015.15091141

12. Muñoz RF, Beardslee WR, Leykin Y. Major depression can be prevented. Am Psychol. 2012;67:285–95. doi: 10.1037/a0027666

13. Miguel C, Amarnath A, Akhtar A, et al. Universal, selective and indicated interventions for supporting mental health at the workplace: an umbrella review of meta-analyses. Occup Environ Med. 2023;80:225–36. doi: 10.1136/oemed-2022-108698

14. Miller AH, Raison CL. The role of inflammation in depression: from evolutionary imperative to modern treatment target. Nat Rev Immunol. 2016;16:22–34. doi: 10.1038/nri.2015.5

15. Dantzer R, O’Connor JC, Freund GG, et al. From inflammation to sickness and depression: when the immune system subjugates the brain. Nat Rev Neurosci. 2008;9:46–56. doi: 10.1038/nrn2297

16. Berk M, Williams LJ, Jacka FN, et al. So depression is an inflammatory disease, but where does the inflammation come from? BMC Med. 2013;11:200. doi: 10.1186/1741-7015-11-200

17. Köhler CA, Freitas TH, Maes M, et al. Peripheral cytokine and chemokine alterations in depression: a meta-analysis of 82 studies. Acta Psychiatr Scand. 2017;135:373–87. doi: 10.1111/acps.12698

18. Pastis I, Santos MG, Paruchuri A. Exploring the role of inflammation in major depressive disorder: beyond the monoamine hypothesis. Front Behav Neurosci. 2023;17:1282242. doi: 10.3389/fnbeh.2023.1282242

19. Roman M, Irwin MR. Novel neuroimmunologic therapeutics in depression: A clinical perspective on what we know so far. Brain Behav Immun. 2020;83:7–21. doi: 10.1016/j.bbi.2019.09.016

20. Arksey H, O’Malley L. Scoping studies: towards a methodological framework. International Journal of Social Research Methodology. 2005;8:19–32. doi: 10.1080/1364557032000119616

21. Levac D, Colquhoun H, O’Brien KK. Scoping studies: advancing the methodology. Implement Sci. 2010;5:69. doi: 10.1186/1748-5908-5-69

22. Tricco AC, Lillie E, Zarin W, et al. PRISMA Extension for Scoping Reviews (PRISMA-ScR): Checklist and Explanation. Ann Intern Med. 2018;169:467–73. doi: 10.7326/M18-0850

23. Nosek BA, Alter G, Banks GC, et al. SCIENTIFIC STANDARDS. Promoting an open research culture. Science. 2015;348:1422–5. doi: 10.1126/science.aab2374

24. Armstrong R, Hall BJ, Doyle J, et al. Cochrane Update. “Scoping the scope” of a cochrane review. J Public Health (Oxf). 2011;33:147–50. doi: 10.1093/pubmed/fdr015

25. Munn Z, Peters MDJ, Stern C, et al. Systematic review or scoping review? Guidance for authors when choosing between a systematic or scoping review approach. BMC Med Res Methodol. 2018;18:143. doi: 10.1186/s12874-018-0611-x

26. Brien SE, Lorenzetti DL, Lewis S, et al. Overview of a formal scoping review on health system report cards. Implement Sci. 2010;5:2. doi: 10.1186/1748-5908-5-2

27. Rumrill PD, Fitzgerald SM, Merchant WR. Using scoping literature reviews as a means of understanding and interpreting existing literature. Work. 2010;35:399–404. doi: 10.3233/WOR-2010-0998

28. Davis K, Drey N, Gould D. What are scoping studies? A review of the nursing literature. Int J Nurs Stud. 2009;46:1386–400. doi: 10.1016/j.ijnurstu.2009.02.010

29. Chen Y, Bo Y. AI-driven early detection of severe influenza in Jiangsu, China: a deep learning model validated through the design of multi-center clinical trials and prospective real-world deployment. Front Public Health. 2025;13:1610244. doi: 10.3389/fpubh.2025.1610244

30. Schardt C, Adams MB, Owens T, et al. Utilization of the PICO framework to improve searching PubMed for clinical questions. BMC Med Inform Decis Mak. 2007;7:16. doi: 10.1186/1472-6947-7-16

31. Diagnostic and statistical manual of mental disorders: DSM-5^TM^, 5th ed. Arlington, VA, US: American Psychiatric Publishing, Inc. 2013.

32. Zheng-Ren Ma L-LM, Fei Zhao YB. Effects of oral calcium on reproduction and postpartum health in cattle: a meta-analysis and quality assessment. Frontiers in Veterinary Science.

33. Liang Q. Effects of home-based telemedicine and mHealth interventions on blood pressure in stroke patients: a systematic evaluation and meta-analysis of randomized controlled trials. Journal of Stroke and Cerebrovascular Diseases. 2024.

34. Bo Y, Zhao F. Platelets as central hubs of inflammation. Front Immunol. 2025;16:1683553. doi: 10.3389/fimmu.2025.1683553

35. Zhao F, Bo Y, Su X-L. Effect of cognitive behavioral therapy on cancer-related fatigue and psychological status in ovarian cancer patients: A meta-analysis. World J Psychiatry. 2025;15:112479. doi: 10.5498/wjp.v15.i12.112479

36. Grey matters: a practical tool for searching health-related grey literature. Canadian Agency for Drugs and Technologies for Health. 2018. https://www.cadth.ca/resources/finding-evidence (accessed 3 January 2025)

37. Grey Matters: A Tool for Searching Health-related Grey Literature. Ottawa: Canada’s Drug Agency. 2024. https://greymatters.cda-amc.ca (accessed 3 January 2025)

38. Moher D, Liberati A, Tetzlaff J, et al. Preferred reporting items for systematic reviews and meta-analyses: the PRISMA statement. PLoS Med. 2009;6:e1000097. doi: 10.1371/journal.pmed.1000097

39. Fergusson D, Monfaredi Z, Pussegoda K, et al. The prevalence of patient engagement in published trials: a systematic review. Res Involv Engagem. 2018;4:17. doi: 10.1186/s40900-018-0099-x

40. Domecq JP, Prutsky G, Elraiyah T, et al. Patient engagement in research: a systematic review. BMC Health Serv Res. 2014;14:89. doi: 10.1186/1472-6963-14-89

41. Zhao F, Guo Z, Bo Y, et al. Is cognitive behavioral therapy an efficacious treatment for psychological interventions in body dysmorphic disorders? A meta-analysis based on current evidence from randomized controlled trials. J Affect Disord. 2024;352:237–49. doi: 10.1016/j.jad.2024.02.004

42. Chen Y, Bo Y, Han Z, et al. Health needs assessment: development of a simplified risk probability scale design for rapid mental health screening in emergency frontline rescue teams. Front Public Health. 2025;13:1601236. doi: 10.3389/fpubh.2025.1601236

43. Staniszewska S, Brett J, Simera I, et al. GRIPP2 reporting checklists: tools to improve reporting of patient and public involvement in research. BMJ. 2017;358:j3453. doi: 10.1136/bmj.j3453

44. Thomas J, Harden A. Methods for the thematic synthesis of qualitative research in systematic reviews. BMC Med Res Methodol. 2008;8:45. doi: 10.1186/1471-2288-8-45

45. Ryan C, Hesselgreaves H, Wu O, et al. Protocol for a systematic review and thematic synthesis of patient experiences of central venous access devices in anti-cancer treatment. Syst Rev. 2018;7:61. doi: 10.1186/s13643-018-0721-x

46. Xiao W, Lian JJ, Ouyang K, et al. Newton downhill optimizer with application to engineering optimization and breast cancer feature selection. Biomedical Signal Processing and Control. 2026;117:109184. doi: 10.1016/j.bspc.2025.109184

47. Zhuang Z, Bo Y. Self-management and role of nurses of diabetic patients: a critical narrative literature review during COVID-19 pandemic. Front Med (Lausanne). 2025;12:1626447. doi: 10.3389/fmed.2025.1626447

