## Supplementary Materials for "The Role of Inflammation in Depression: A Scoping Review Protocol in Mechanisms, Evidence, and Therapeutic Potential"

### 1 Search Formula

#### 1.1 PubMed (2025-3-12: 1575 hits); (2025-3-16: 1577 hits); (2025-3-17: 924 hits)

1#: ((((((((((((((((((((((((((((Depressive Disorder) OR (Depressive Disorders)) OR (Disorder, Depressive)) OR (Disorders, Depressive)) OR (Neurosis, Depressive)) OR (Depressive Neuroses)) OR (Depressive Neurosis)) OR (Neuroses, Depressive)) OR (Depression, Endogenous)) OR (Depressions, Endogenous)) OR (Endogenous Depression)) OR (Endogenous Depressions)) OR (Melancholia)) OR (Melancholias)) OR (Unipolar Depression)) OR (Depression, Unipolar)) OR (Depressions, Unipolar)) OR (Unipolar Depressions)) OR (Depressive Syndrome)) OR (Depressive Syndromes)) OR (Syndrome, Depressive)) OR (Syndromes, Depressive)) OR (Depression, Neurotic)) OR (Depressions, Neurotic)) OR (Neurotic Depression)) OR (Neurotic Depressions)) OR ("Depressive Disorder"[Mesh])) OR ((((((Depression) OR (Depressive Symptoms)) OR (Depressive Symptom)) OR (Symptom, Depressive)) OR (Emotional Depression)) OR (Depression, Emotional))) OR ("Depression"[Mesh])

2#: ((("Inflammation"[Mesh]) OR (((((Inflammation) OR (Inflammations)) OR (Innate Inflammatory Response)) OR (Inflammatory Response, Innate)) OR (Innate Inflammatory Responses))) OR ("Neurogenic Inflammation"[Mesh])) OR ((((Neurogenic Inflammation) OR (Inflammation, Neurogenic)) OR (Inflammations, Neurogenic)) OR (Neurogenic Inflammations))

3#: 1# AND 2#

#### 1.2 Web Of Science (2025-3-12: 4294 hits); (2025-3-17: 122 hits)

1#: TS=( Depressive Disorder OR Depressive Disorders OR Depressive Neuroses OR  Depressive Neurosis OR Endogenous Depression OR Endogenous Depressions OR Melancholia OR Melancholias OR Unipolar Depression OR Unipolar Depressions OR Depressive Syndrome OR Depressive Syndromes OR Neurotic Depression OR Neurotic Depressions OR Depression OR Depressive Symptoms OR Depressive Symptom OR Emotional Depression )

2#: TS=( Inflammation OR Inflammations OR Innate Inflammatory Response OR  Innate Inflammatory Responses OR Neurogenic Inflammation OR Neurogenic Inflammation OR Neurogenic Inflammation OR Neurogenic Inflammations )

3#: 1# AND 2#

#### 1.3 Scopus (2025-3-12: 5031 hits); (2025-3-17: 74 hits)

#1: TITLE-ABS-KEY( "Depressive Disorder" OR "Depressive Disorders" OR "Depressive Neuroses" OR "Depressive Neurosis" OR "Endogenous Depression" OR "Endogenous Depressions" OR "Melancholia" OR "Melancholias" OR "Unipolar Depression" OR "Unipolar Depressions" OR "Depressive Syndrome" OR "Depressive Syndromes" OR "Neurotic Depression" OR "Neurotic Depressions" OR "Depression" OR "Depressive Symptoms" OR "Depressive Symptom" OR "Emotional Depression" )

#2: TITLE-ABS-KEY( "Inflammation" OR "Inflammations" OR "Innate Inflammatory Response" OR "Innate Inflammatory Responses" OR "Neurogenic Inflammation" OR "Neurogenic Inflammation" OR "Neurogenic Inflammation" OR "Neurogenic Inflammations" )

#3: #1 AND #2

#### 1.4 Cochrane (2025-3-12: 2276 hits);(2025-3-16: 2211 hits)

#1: MeSH descriptor: [Depressive Disorder] explode all trees
#2: MeSH descriptor: [Depression] explode all trees
#3: ("Depressive Disorder" OR "Depressive Disorders" OR "Depressive Neuroses" OR "Depressive Neurosis" OR "Endogenous Depression" OR "Endogenous Depressions" OR "Melancholia" OR "Melancholias" OR "Unipolar Depression" OR "Unipolar Depressions" OR "Depressive Syndrome" OR "Depressive Syndromes" OR "Neurotic Depression" OR "Neurotic Depressions" OR "Depression" OR "Depressive Symptoms" OR "Depressive Symptom" OR "Emotional Depression"):ti,ab,kw
#4: MeSH descriptor: [Inflammation] explode all trees
#5: MeSH descriptor: [Neurogenic Inflammation] explode all trees
#6: ("Inflammation" OR "Inflammations" OR "Innate Inflammatory Response" OR "Innate Inflammatory Responses" OR "Neurogenic Inflammation" OR "Neurogenic Inflammations"):ti,ab,kw
#7: (#1 OR #2 OR #3) AND (#4 OR #5 OR #6)

#### 1.5 Embase (2025-3-12: 2421 hits); (2025-3-16: 12735 hits) ; (2025-3-17: 1230 hits)

#1: **'depressive disorder'**/exp OR **'depressive disorder'** OR **'depression'**/exp OR **'depression'**
#2: **'depressive disorder'**:ti,ab,kw OR **'depressive disorders'**:ti,ab,kw OR **'depressive neuroses'**:ti,ab,kw OR **'depressive neurosis'**:ti,ab,kw OR **'endogenous depression'**:ti,ab,kw OR **'endogenous depressions'**:ti,ab,kw OR **'melancholia'**:ti,ab,kw OR **'melancholias'**:ti,ab,kw OR **'unipolar depression'**:ti,ab,kw OR **'unipolar depressions'**:ti,ab,kw OR **'depressive syndrome'**:ti,ab,kw OR **'depressive syndromes'**:ti,ab,kw OR **'neurotic depression'**:ti,ab,kw

OR **'neurotic depressions'**:ti,ab,kw OR **'depression'**:ti,ab,kw OR **'depressive symptoms'**:ti,ab,kw OR **'depressive symptom'**:ti,ab,kw OR **'emotional depression'**:ti,ab,kw
#3: **'inflammation'**/exp OR **'neurogenic inflammation'**/exp
#4: **'inflammation'**:ti,ab,kw OR **'inflammations'**:ti,ab,kw OR **'innate inflammatory response'**:ti,ab,kw OR **'innate inflammatory responses'**:ti,ab,kw OR **'neurogenic inflammation'**:ti,ab,kw OR **'neurogenic inflammations'**:ti,ab,kw
#5: (#1 OR #2) AND (#3 OR #4)

#### 1.6 arXiv [https://arxiv.org]

("Depression" OR "Depressive Disorder" OR "Depressive Disorders" OR "Neurotic Depression" OR "Unipolar Depression" OR "Endogenous Depression" OR "Melancholia" OR "Depressive Neuroses" OR "Depression, Emotional" OR "Depressive Syndrome" OR "Depressive Symptoms")
AND
("Inflammation" OR "Inflammations" OR "Innate Inflammatory Response" OR "Inflammatory Response" OR "Neurogenic Inflammation" OR "Neurogenic Inflammations")

#### 1.7 BioRxiv [https://www.biorxiv.org]

("Depression" OR "Depressive Disorder" OR "Unipolar Depression" OR "Neurotic Depression" OR "Melancholia" OR "Endogenous Depression" OR "Depressive Symptoms" OR "Depressive Syndrome")
AND
("Inflammation" OR "Innate Inflammatory Response" OR "Inflammatory Response" OR "Neurogenic Inflammation")

#### 1.8 F1000 [https://f1000research.com]

("Depression" OR "Depressive Disorder" OR "Depressive Symptoms" OR "Endogenous Depression" OR "Unipolar Depression")
AND
("Inflammation" OR "Neurogenic Inflammation" OR "Inflammatory Response" OR "Innate Inflammatory Response")

#### 1.9 FigShare [https://figshare.com]

("Depression" OR "Depressive Disorder" OR "Unipolar Depression" OR "Melancholia")
AND
("Inflammation" OR "Innate Inflammatory Response" OR "Neurogenic Inflammation")

#### 1.10 PeerJ Preprints [https://peerj.com/preprints]

("Depression" OR "Depressive Disorder" OR "Neurotic Depression" OR "Depression, Emotional")
AND
("Inflammation" OR "Neurogenic Inflammation" OR "Innate Inflammatory Response")

#### 1.11 Zenodo [https://zenodo.org]

("Depression" OR "Depressive Disorders" OR "Unipolar Depression" OR "Depressive Symptoms")
AND
("Inflammation" OR "Neurogenic Inflammation" OR "Inflammatory Response")

#### 1.12 Oxford University ePrints [https://eprints.ox.ac.uk]

("Depression" OR "Depressive Disorder" OR "Endogenous Depression" OR "Depressive Symptoms" OR "Unipolar Depression")
AND
("Inflammation" OR "Neurogenic Inflammation" OR "Innate Inflammatory Response")

#### 1.13 Nature Precedings [https://www.nature.com/precedings]

("Depression" OR "Depressive Disorder" OR "Unipolar Depression" OR "Neurotic Depression")
AND
("Inflammation" OR "Neurogenic Inflammation" OR "Inflammatory Response")

#### 1.14 medRxiv [https://www.medrxiv.org]

("Depression" OR "Depressive Disorder" OR "Endogenous Depression" OR "Depressive Symptoms")
AND
("Inflammation" OR "Neurogenic Inflammation" OR "Innate Inflammatory Response")

#### 1.15 Peerage of Science [https://www.peerageofscience.org]

("Depression" OR "Depressive Disorder" OR "Unipolar Depression" OR "Melancholia")
AND
("Inflammation" OR "Innate Inflammatory Response" OR "Neurogenic Inflammation")
